# ROBIN: A unified nanopore-based sequencing assay integrating real-time, intraoperative methylome classification and next-day comprehensive molecular brain tumour profiling for ultra-rapid tumour diagnostics

**DOI:** 10.1101/2024.09.10.24313398

**Authors:** S Deacon, I Cahyani, N Holmes, G Fox, R Munro, S Wibowo, T Murray, H Mason, M Housley, D Martin, A Sharif, A Patel, R Goldspring, S Brandner, F Sahm, S Smith, SML Paine, M Loose

## Abstract

**Background:** Advances in our technological capacity to interrogate brain tumour biology has led to the ever-increasing use of genomic sequencing in routine diagnostic decision making. Presently, brain tumours are routinely classified based on their epigenetic signatures, leading to a paradigm shift in diagnostic pathways. Such testing can be performed so rapidly using nanopore sequencing that results can be provided intraoperatively. This information greatly improves upon the fidelity of smear diagnosis and can help surgeons tailor their approach, balancing the risks of surgery with the likely benefit. Nevertheless, full integrated diagnosis may require subsequent additional assays to detect pathognomonic somatic mutations and structural variants, thereby delaying the time to final diagnosis.

**Methods:** Here, we present ROBIN, a tool based upon PromethION nanopore sequencing technology that can provide both real-time, intraoperative methylome classification and next-day comprehensive molecular profiling within a single assay. ROBIN uniquely integrates three methylation classifiers ^1–3^ to improve diagnostic performance in the intraoperative setting.

**Findings:** We demonstrate classifier performance on 50 prospective intraoperative cases, achieving a diagnostic turnaround time under 2 hours and generating robust tumour classifications within minutes of sequencing. Furthermore, ROBIN can detect single nucleotide variants (SNVs), copy number variants (CNVs) and structural variants (SVs) in real-time, and is able to inform a complete integrated diagnosis within 24 hours. Classifier performance demonstrated concordance with final integrated diagnosis in 90% of prospective cases.

**Interpretation:** Nanopore sequencing can greatly improve upon the turnaround times for standard of care diagnostic testing, including sequencing, and is furthermore able to reliably provide clinically actionable intraoperative tumour classification.

**Funding:** The Jean-Shanks Foundation, the Pathological Society of Great Britain and Ireland, the British Neuropathological Society, and the Wellcome Trust.

## Box 1

Research in context

## Evidence before this study

The 2021 WHO classification of tumours of the central nervous system mandated the use of molecular testing in the diagnosis of brain tumours. Whilst various molecular features now define tumour subtypes, the recent understanding that pan-genome CpG methylation profiling can robustly delineate tumour subtypes has demonstrated particular utility and is currently revolutionising brain tumour diagnostic pathways. We searched PubMed from its inception to 1^st^ September 2024 for articles relevant to nanopore-based classification of brain tumours (i.e. using the search terms “nanopore” AND “classification” AND “tumour”). Additionally, articles were searched for relevant citations. Currently, the present clinical implementations of such methylation testing are predicated on the use of array-based technologies, where requisite sample transport and batch processing prolongs turnaround times, with published times of approximately three weeks. As such, a key challenge of delivering expedient cancer diagnostics is integrating genomic testing into standard of care pathways such that results can be provided to the clinical team within a clinically relevant timeframe.

## Added value of this study

A key challenge for intraoperative tumour classification is balancing the competing needs of rapid sample processing and the requirement to generate DNA of adequate yield and read length for the subsequent detection of additional molecular features. Previous implementations of intraoperative, ultra-fast classification do not generate libraries of sufficient read length and pore occupancy for detecting additional molecular features beyond methylation-based classification alone. Implementations of nanopore sequencing for such comprehensive diagnostics have previously relied on ligation-based approaches that are not suitable for intraoperative use due to their longer turnaround time. To our knowledge, our study is the first to demonstrate an ultra-fast workflow which can integrate intraoperative classification and next-day full molecular profiling within a single assay. Furthermore, our study builds upon previous work by integrating three analytic tools to generate a robust intraoperative classification and improve diagnostic confidence, in contrast to previous work which rely on a single classification alone.

## Implications of all the available evidence

This assay greatly improves upon SoC diagnostic turnaround times and offers the opportunity to tailor surgical approach to the individual patient and initiate adjuvant treatment earlier. While currently limited to brain tumour diagnostics, this technology serves as the harbinger of near-patient, ultra-rapid pan-cancer tumour sequencing and is well situated to revolutionise oncological care in a wide range of cancers.

## Background

The 2021 WHO classification of CNS tumours substantially increased the mandated use of molecular testing in the diagnosis of brain tumours and, in particular, DNA methylation-based classification is now an essential criterion in the routine diagnosis of many tumour subtypes ^4^. DNA methylation of cytosines is maintained across cell divisions and is a means by which histogenesis can be traced to the cellular origin, despite neoplastic dedifferentiation ^5,6^. Currently, testing is implemented by microarray-based methods, such as Illumina EPIC arrays which, because of high capital costs, are restricted to specialist tertiary care centres where high-throughput can reduce per-assay costs ^7^. This requires inter-regional sample transport and multiplexed batch processing, which can cause diagnostic delays, with average turnaround times between several days to weeks ^7^.

In contrast to array-based technologies and unravelling of a comprehensive molecular diagnosis by multiple assays (such as Methylome classification including copy number assay, DNA NGS and RNA NGS), nanopore sequencing (Oxford Nanopore Technologies) enables assessment of copy-number profile, mutational and methylation analysis in a single assay via native strand, long-read sequencing. Capital and consumable costs are relatively low, and analysis can be performed on individual samples. Nanopore sequencing has been shown to confidently classify brain tumours based on methylation status ^1,8^. Crucially, this technology enables affordable local sequencing with much faster turn-around times compared to the current standard of care (SoC) ^2,9,10^.

Current implementations of nanopore-based classification typically rely on low-coverage approaches using MinION flow cells, sequencing a small fraction of the genome’s total CpG sites. At such an ultra-low read depth, only binary methylation information can be obtained at most sites, and specialised tools have been developed to classify accurately based on this sparse methylation data ^2^. However, these tools are limited and are not able to report additional data such as SNVs and SVs with high confidence due to low coverages. Nevertheless, capturing such genetic alterations is required to form a WHO integrated diagnosis, and so additional assays such as RNA and DNA sequencing are required.

Here, we present an improved analysis pipeline performing integrated methylation-based tumour classification, copy number profiling, structural variant calling, *MGMT* promoter methylation, and mutation detection. We use higher-coverage PromethION flow cells and a modified transposase-based library preparation. We demonstrate the feasibility of using this technique for both intraoperative tumour classification and next-day final molecular diagnosis.

## Methods

### Patient selection

Thirty cases with primary CNS tumour and available adequate frozen tissue were selected for retrospective sequencing as per previously published protocols (supplementary materials)^8^. For the prospective cohort, patients were selected for nanopore-based sequencing if anticipated to have a primary intracranial neoplasm and be eligible for SoC methylation array. Consent for nanopore sequencing was obtained by the neurosurgical team preoperatively. However, four patients included in the study did not warrant further molecular testing by SoC approaches: one WHO grade 1 meningioma with typical meningothelial morphology and no concerning features, and two classical glioblastomas with no *IDH1* mutation by immunohistochemistry in patients over the age of 60 years, and a germinoma with diagnostic immunohistochemistry. As such, for these cases the nanopore classifications were compared against the final WHO Integrated Diagnosis alone, although nanopore-sequencing nevertheless provided rapid evaluation of *MGMT* promoter methylation in the cases of glioblastoma. Once case was excluded from the study during intraoperative DNA extraction due to the smear demonstrating metastatic adenocarcinoma.

### Intraoperative DNA extraction

Tissue was collected fresh, direct from the operating theatre and three spatially distant tumour samples were selected for further processing. All tissue selected was in excess of that required for SoC neuropathological workup. For each sample, between 5 mg - 25 mg tissue was taken for DNA extraction, and adjacent tissue was reserved for intraoperative smear. DNA extraction was performed using the QIAmp Fast DNA Tissue Kit, as per the manufacturer’s instructions. During the DNA extraction, the neuropathologist reported in parallel on the smear preparation and advised which of the three samples was most suitable for sequencing, for example excluding samples with extensive necrosis or a low tumour cell fraction. All DNA extraction reagents are stable at room temperature, avoiding the need for specialist storage and enabling the preparation of reagents prior to the operation, enabling immediate tissue processing upon their receipt.

### Intraoperative library preparation and sequencing

Samples were prepared with the ONT ultra-long kit using an adjusted protocol (https://protocols.io/view/intra-operative-nanopore-sequencing-to-classify-br-c65qzg5w). Briefly, 600 ng input DNA was mechanically sheared and tagmented. DNA was then purified using the AmpureXP DNA binding beads (Beckman Coulter; A63882). Library concentration was measured using the Invitrogen™ Qubit™ 3 Fluorometer and High Sensitivity assay after shearing and prior to flow cell loading. The library was loaded onto R10.4.1 PromethION flowcells. Sequencing was commenced five minutes after library loading and performed for up to 24 hours. Basecalling was performed whilst sequencing using MinKNOW. Reads were called using the high accuracy model with 5mC and 5hmC modifications. Reads were mapped to GRCh38 during basecalling and the resulting BAM files used for subsequent analysis. Readfish was used for adaptive sequencing (supplementary materials)^11^.

### Real-time intraoperative visualisation and reporting

To facilitate real-time analysis of data we have developed ROBIN (Rapid nanpOre Brain intraoperatIve classificatioN). ROBIN provides a graphical interface to present sequencing results as data are being generated (https://github.com/looselab/robin). ROBIN integrates multiple classifiers of methylation, copy number changes, coverage over targets and single nucleotide variants, *MGMT* promoter methylation and indicates candidate fusion genes during sequencing. Methylation classifiers include Sturgeon, CrossNN and the random forest approach from RapidCNS2 ^2,3,8^. Copy number changes are dynamically visualised using CNV from BAM (https://github.com/adoni5/cnv_from_bam). Single nucleotide variants are analysed using ClairS and Clair-To (https://github.com/HKU-BAL/ClairS). Resultant variants are annotated using snpEff and snpSift ^12,13^. *MGMT* methylation status is analysed as in RapidCNS2 and visualised using methylartist ^8,14^. Candidate gene fusions are identified by inspection of long reads linking two targets in the adaptive sampling BED file or linking one gene from the BED file and one target elsewhere in the genome. Fusions are visualised using https://github.com/Edinburgh-Genome-Foundry/DnaFeaturesViewer. ROBIN operates through a web interface, although running on the local computer performing the sequencing. As a consequence, the output of ROBIN can be viewed by a pathologist remote to the sequencing computer, enabling an integrated diagnosis by a clinician in real-time during sequencing. Whilst the results of nanopore-based tumour sequencing and classification were available prior to the completion of SoC testing, this information was treated strictly as research use only and no data were used to inform patient care.

## Results

### Same-day methylation-based CNS tumour classification

Nanopore sequencing enables rapid methylation-based tumour classification. Thirty frozen samples were retrospectively sequenced using MinION flow cells to validate the methodology, and fifty tumours were subsequently prospectively sequenced intraoperatively using PromethION flow cells. Of these, 24 (80%) and 45 (90%) of the respective cohorts were correctly classified by nanopore after 24 hours total sequencing time (figure 1). Classification results were validated against both SoC methylation array and final integrated diagnosis (supplementary data tables 1 and 2). The major causes of misclassification were common across both cohorts (supplementary data tables 3 and 4), including misclassifications of novel entities absent from the nanopore classifiers and low tumoral DNA in sample tissue. In neither the retrospective nor prospective cohorts, did the ONT-based pipeline deliver an erroneous result the reasons for which could not be understood/accounted for by either sample bias or differences in classifier versions.

**Figure 1:**
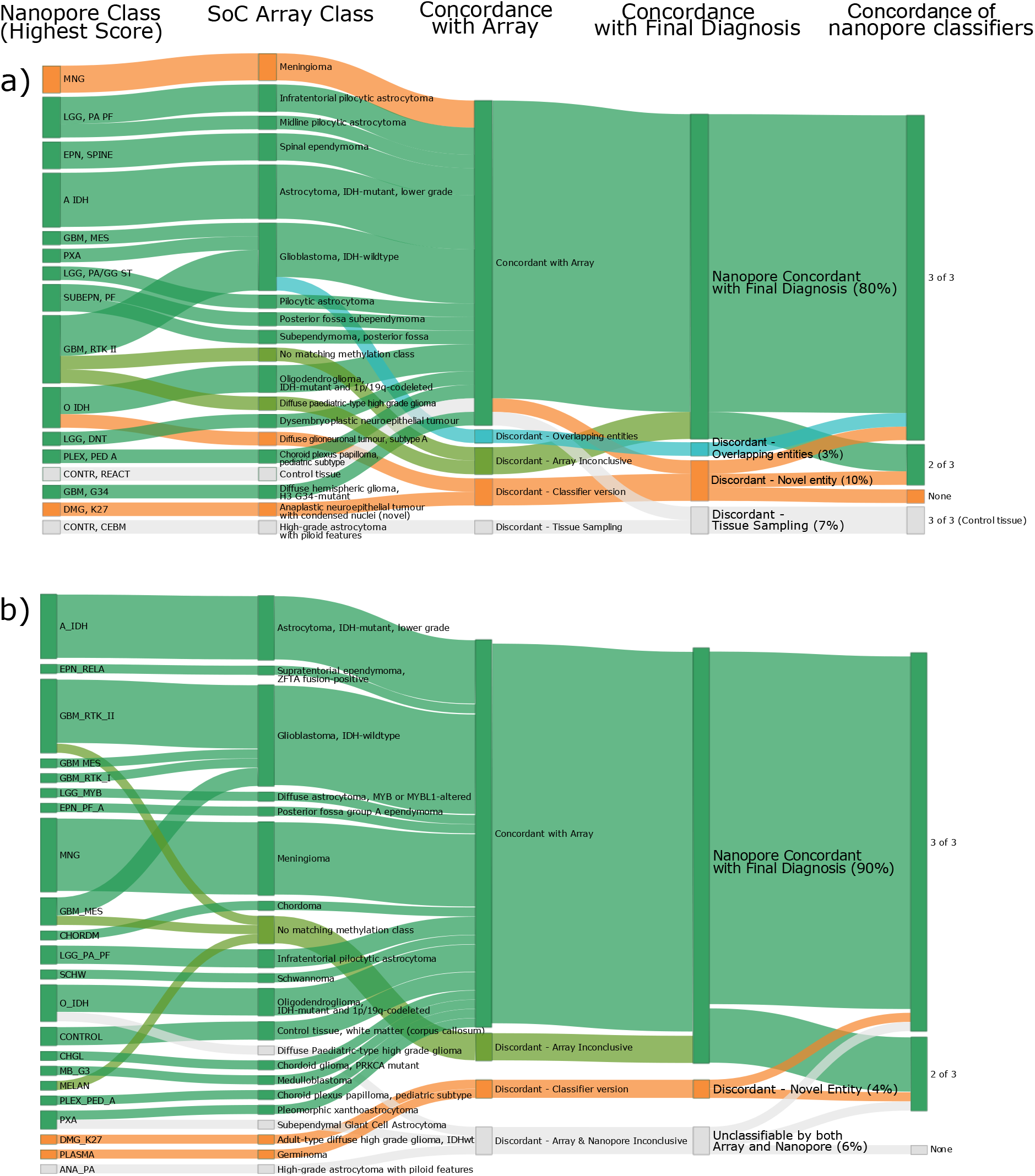
Nanopore classifications versus SoC testing. A) Retrospective cohort. B) Prospective (intraoperative cohort). Highest-scoring nanopore class after 24 hours is displayed, excepting a case of chordoma which was correctly classified intraoperatively yet classified as control tissue after 24 hours. Concordance with array was determined irrespective of glioblastoma subclass.

### Next-day comprehensive molecular profiling of CNS tumours

Alongside methylation-based classification, other molecular features are crucial for accurate diagnosis. For example, copy number changes are part of some WHO diagnostic criteria (figure 2) ^4^. Since both on-target and off-target reads contribute to the copy number data, these plots can be rapidly and intraoperatively generated by ROBIN, with diagnostic evidence for copy number changes apparent even at low coverages. Copy number profiles generated by ROBIN were in concordance with array-generated data. For example, most cases of glioblastoma demonstrated the canonical gain of chromosome 7 and loss of chromosome 10, and all cases of oligodendroglioma exhibited the pathognomonic loss of chromosomes 1p and 19q (supplementary data figure 1). Additionally, gene-level characteristics such as co-deletion of CDKN2A/B in astrocytoma are reliably detected (supplementary data figure 2).

**Figure 2:**
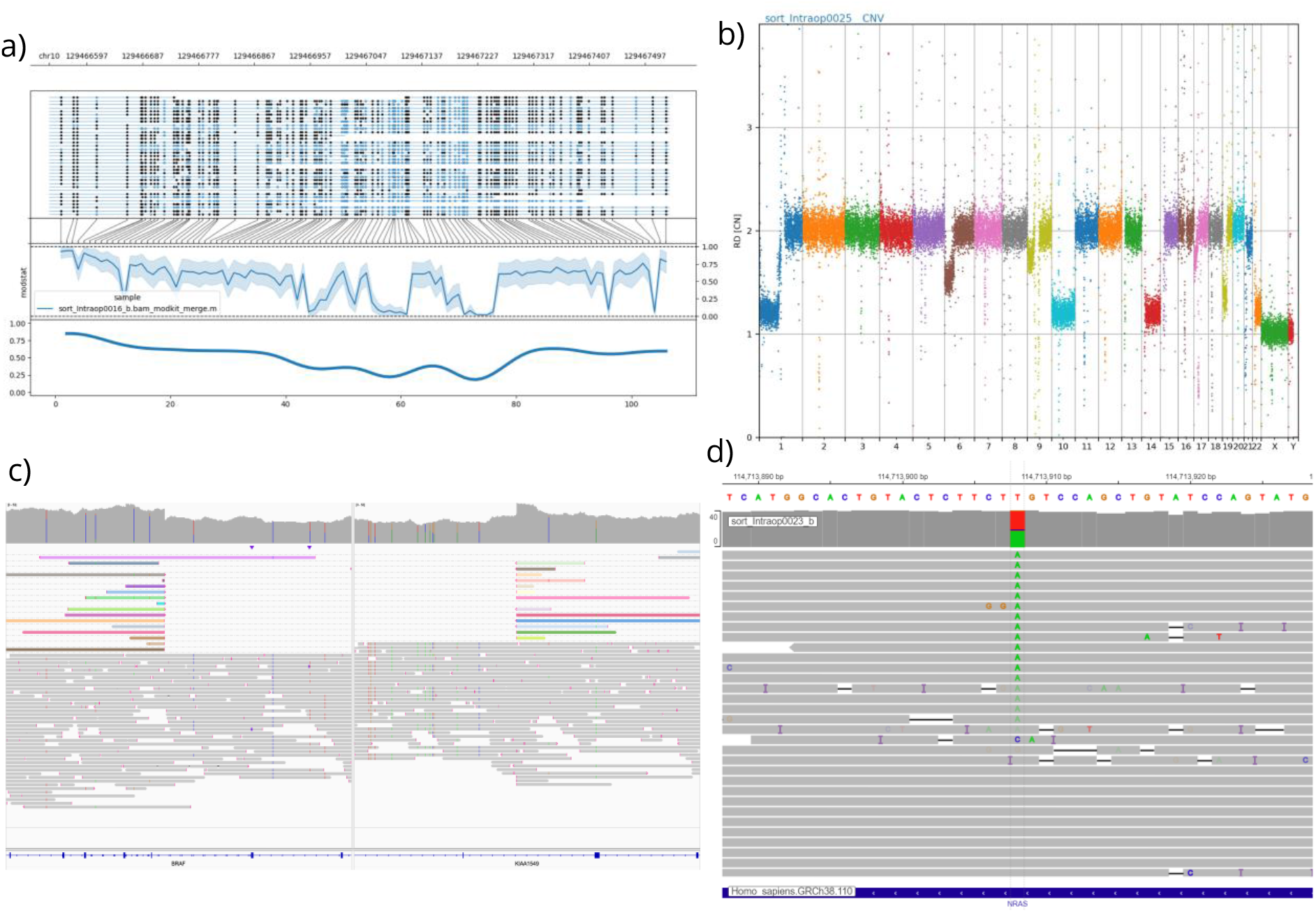
Additional molecular features captured within the next-day report. A) Methylation distribution at the MGMT promoter in a glioblastoma. Top pane: each line corresponds to a single read; black dots indicate methylated sites and blue dots indicate non-methylated sites. Middle pane: average methylation score across the MGMT promoter. Bottom pane: smoothed average. Clinically relevant methylation is defined as >25% methylation across the region. B) Copy number plot of an oligodendroglioma, demonstrating the canonical codeletion of 1p and 19q chromosome arms. C) Demonstration of a pathognomonic BRAF::KIAA1549 fusion in a pilocytic astrocytoma. D) Demonstration of a c.A182T (p.Q61L) mutation of NRAS in a case of metastatic melanoma.

A key strength of adaptive sampling coupled with PromethION flow cells is that much higher coverage can be generated, within a shorter timeframe when compared to standard MinION-based approaches. This enabled confident identification of relevant SNVs and SVs in our cohort, evaluated against known tumour hallmarks and, where available, SoC testing. *IDH1/*2 mutations were correctly identified in 3 of 3 oligodendrogliomas and 6 of 7 astrocytomas; in one case low tumour DNA fraction impaired analysis (supplementary data table 6). Identification of specific structural variants is critical in the diagnosis of certain tumours, and we were able to identify relevant fusions in 6 cases (supplementary data figure 3). The distinct advantage of long-read sequencing is demonstrated by the identification of novel, complex fusions in cases such as a MYB-altered astrocytoma and an VGLL-fused intracranial schwannoma.

The methylation level of the *MGMT* promoter is a clinically important biomarker, identifying patients who are likely to respond to adjuvant temozolomide chemotherapy. Nanopore-derived *MGMT* status was concordant with array-based predicted *MGMT* status in 44 of 46 cases in the prospective cohort (supplementary data table 7). One discrepant case had a methylation score very close to the cut-off score, suggesting potentially a subtle difference in methylation thresholds between array and nanopore approaches. Indeed, such equivocal low-level methylation of *MGMT* may still be an indicator of potential sensitivity to temozolomide treatment ^15^.

### Intraoperative methylation-based classification

A key challenge for intraoperative tumour classification is balancing the competing needs of rapid sample processing and the requirement to generate DNA of adequate yield and read length for the subsequent detection of additional molecular features. Previous implementations of nanopore-based comprehensive diagnosis, beyond methylation classification alone, have used ligation approaches and are not suitable for intraoperative use due to their longer turnaround time ^8^. Similarly, whilst our retrospective cohort generated libraries of sufficient quality, overnight tissue lysis and a ligation-based library inordinately delayed time to diagnosis. Our library protocol utilised an adapted transposase-based approach that enabled rapid sample preparation within 90 minutes of tissue receipt, yet preserved an optimal DNA read length for adaptive sampling (figure 3a, 3b).

**Figure 3:**
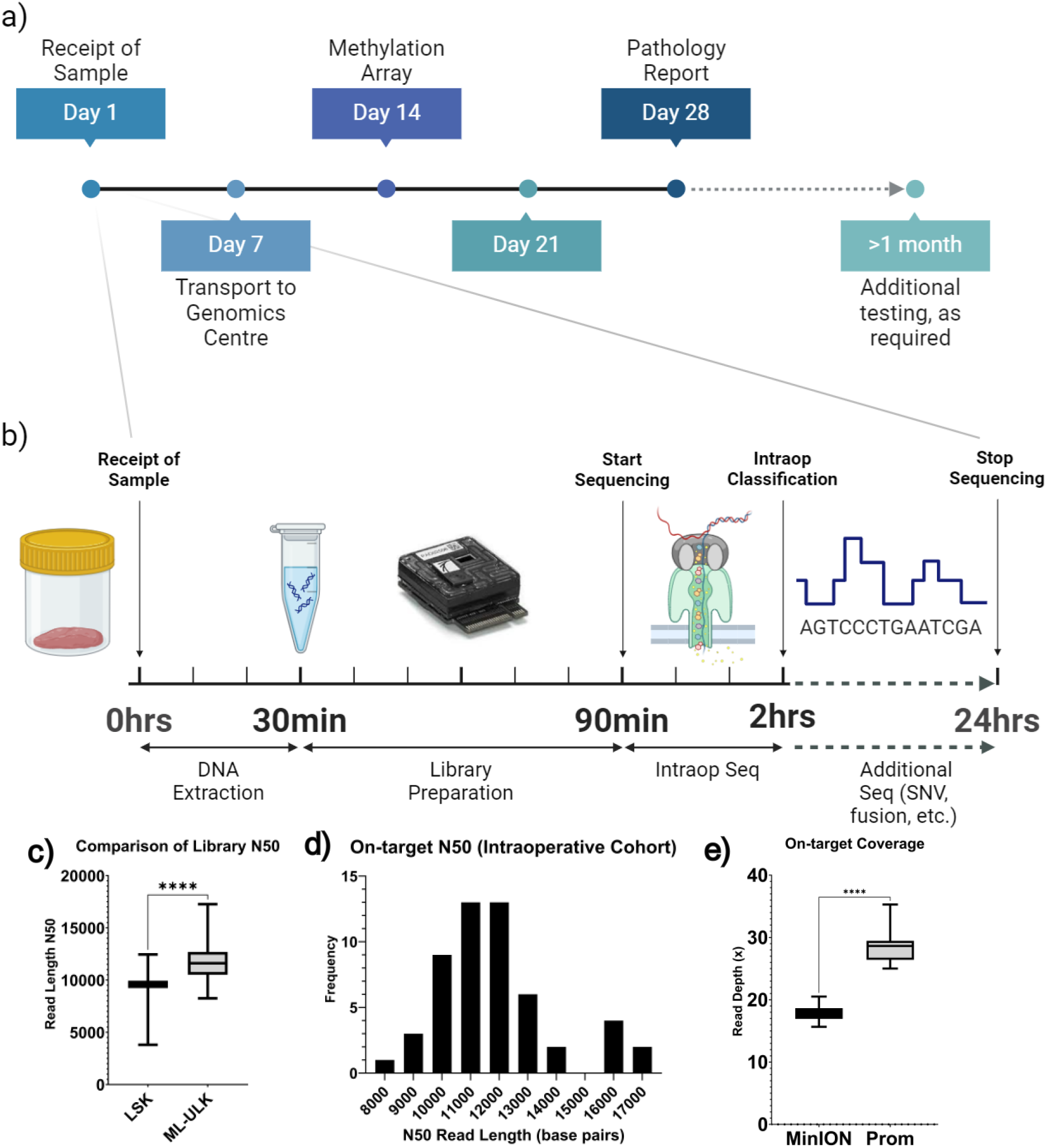
Intraoperative library preparation and sequencing workflow. A) Typical timeline of SoC testing. B) Exemplar intraoperative DNA extraction and sequencing workflow. C) Average library N50 of the intraoperative protocol improved upon that of the retrospective cohort. D) Histogram of on-target N50 values for the intraoperative cohort (mean N50 = 11,739bp). E) On-target coverage was significantly superior in the intraoperative cohort cases (PromethION; runtime 24hrs) compared to the retrospective validation cohort (MinION; runtime 72hours).

Conversely, ultra-fast approaches that require highly disruptive mechanical lysis steps and obviate the need for DNA purification steps do not generate libraries of sufficient read length and pore occupancy for detecting additional molecular features beyond methylation-based classification ^2^. Indeed, whilst these authors opted not to utilise adaptive sampling in the intraoperative setting, it is precisely these additional molecular features, such as SNV and fusion calling, which most benefit from such enrichment, if used alongside libraries of appropriate quality. In our data, our intraoperative protocol produced longer read length libraries compared to the retrospective cohort which followed a previously published ligation-based protocol using g-TUBE shearing ^8^ (figure 3c). For the intraoperative cohort, we achieve a mean N50 of 11,739bp, compared to 9195bp of the ligation-based protocol (*p*<0.0001) (figure 3d), and a significantly higher on-target coverage using PromethION flowcells (figure 3e).

Thirty-eight cases (76%) within the prospective cohort were classified intraoperatively, defined as a confident classification by two or more classifiers within 1 hour of sequencing. Classifiers demonstrated variable performance intraoperatively (supplementary data figure 4). Both neural network-based classifiers (Sturgeon and CrossNN) were able to confidently classify cases within minutes, whereas random forest performance was slower (figure 4a). In a subset of cases, classifier performance was poor during early intraoperative sequencing, due to the sparsity of the data and overall low coverage of CpG sites. The Sturgeon classifier demonstrated a tendency to over-confidently misclassify cases, dynamically switching class mid-sequencing (figure 4b). As such, multiple classifiers are particularly important during intraoperative classification, where congruency of classification can aid clinical interpretation in such cases, or where a single tool fails to classify (figure 4c).

**Figure 4:**
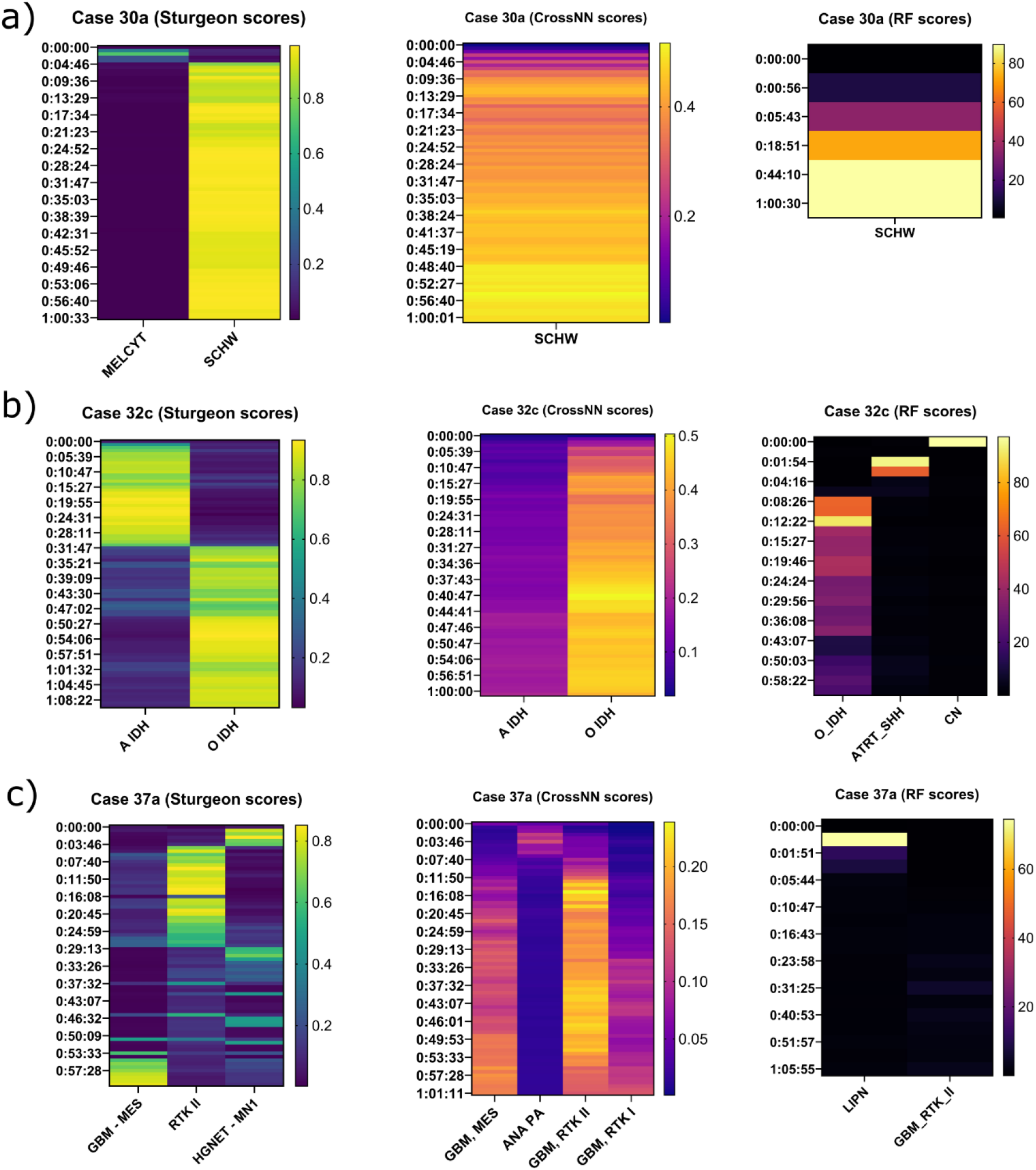
Classifiers confidently predict tumour type during intraoperative sequencing. A) Case example of intracerebral schwannoma which was confidently classified on all three classifiers. B) Example of a case of Oligodendroglioma in which there was some heterogeneity between- and within-classifiers during intraoperative sequencing, but ultimately all classifiers reached concordance. C) Example of a case in which one classifier failed to confidently predict the tumour subtype, yet the remaining two classifiers were successful.

## Discussion

We present a protocol that can generate a comprehensive molecular diagnosis within an improved timeframe compared with current SoC diagnosis. In our prospective cohort, the mean turnaround time for SoC methylation-based classification was 33 days (range: 19 to 81 days). This turnaround time encompasses both delays locally, such as neuropathological assessment and immunohistochemical studies prior to referral (mean: 12 days), and delays following receipt of the sample at the specialist laboratory (mean: 21 days). Whilst local delays may be mitigated by routinely requesting methylation array analysis prior to initial neuropathological workup, such an approach may limit the selection of optimal tissue for molecular studies and risks costly array testing in cases where it is either unsuitable or not required for diagnosis. In the subset of cases where further molecular testing such as RNA- and DNA-based next-generation sequencing was required for SNV and fusion-calling, the mean turnaround time was 68 days (range: 54 to 103 days).

The equivalent mean turnaround times for our ultrafast pipeline were within 24 hours for final comprehensive diagnosis of the 90% of cases which were classifiable. In many cases, methylation classification and identification of relevant SNVs and CNVs were achieved within a few hours of sequencing, but precise quantification of time-to-result is complex. Implementing nanopore-based sequencing for routine practice would democratise access to rapid results, with immediate benefits to patients, medical professionals and the wider health system. Nanopore-based intraoperative classification enables characterisation of the tumour subtype in far greater detail than smear or frozen section alone. Expert neuropathological examination of the smear, whilst undeniably a cornerstone of intraoperative diagnosis, will never attain to the degree of nuance offered by such sequencing; not least because many tumour entities are now explicitly defined by their molecular hallmarks in the absence of defining morphological features.

Despite being of a modest size from a single UK centre, our prospective cohort nevertheless well illustrates the complexity of neuropathological diagnoses and the potential impact of this technology on clinical practice. For example, many types of paediatric tumour consist of non-specific, primitive cells on intraoperative smear and have a wide differential diagnosis. In one paediatric case, the surgical decision was made for subtotal resection awaiting full molecular diagnostic workup. The child subsequently required repeat surgery once SoC testing had been performed and diagnosed the tumour as a RELA::ZFTA-fused ependymoma after 19 days postoperatively. In contrast, nanopore sequencing applied to this sample correctly classified the tumour intraoperatively; a diagnosis which could have spared the patient unnecessary surgical risk. Rare tumours are also often diagnostically challenging, and this may delay the initiation of appropriate treatment or risk over-treatment. In our cohort, we rapidly classified an intracranial schwannoma within minutes of sequencing and demonstrated a novel pathognomonic VGLL3 fusion the following day. Preoperative imaging and clinical assessment were suggestive of an IDH-mutant astrocytoma. Astrocytoma are CNS WHO grades 2 or 3 and confer a lifelong risk of recurrence and high-grade transformation, whereas Schwannoma is CNS WHO grade 1 and surgery is curative. In this context, rapid nanopore diagnosis could have spared the patient weeks of anxiety and distress waiting for a comprehensive diagnosis. Another major strength of nanopore-based classification is its minimal input tissue requirement, making classification of such stereotactic biopsies feasible.

Despite these advantages, neuropathologist interpretation and integration of the data remains crucial. During sequencing, the pathologist can situate the real-time results within the context of the smear examination and the broader clinical and radiological differential diagnosis, ensuring that methylation classification is congruent with these data. For example, some cases which classified as control tissue by one classifier were successfully classified as astrocytoma on the remaining two classifiers. Caution must be taken in interpreting such results; a classification of tumour in two of three classifiers may be accepted by the neuropathologist if, for example, these findings are consistent with the intraoperative smear. Conversely, in our cohort, nanopore sequencing was able to correctly classify a case of melanoma that was misdiagnosed on intraoperative smear. If in keeping with the clinical context, such data may prompt the neuropathologist to re-examine the smear, request more representative tissue in order to perform a second intraoperative smear or expedite adjuvant tests. As analysis is rapid, if the pipeline does not produce diagnostically useful data and additional material is available, DNA can be extracted from a different region and analysed with only a minimal delay.

A definitive, quantitative threshold for classification is challenging to define. We propose that the classification scores must be considered within the clinical context, and that the concordance between multiple classifiers may generate stronger evidence of a robust classification. In our data, using multiple classifiers is vital to inform intraoperative interpretation, thus reducing the risk of misdiagnosis based upon a single datapoint whilst potentially enabling a more definitive diagnosis in indeterminate cases. Similarly, the clinical relevance of copy number changes is highly variable, and the neuropathologist is again vital to the accurate interpretation of these changes on a case-by-case basis. For example, all *IDH*-mutant astrocytoma in our cohort classified as low grade by methylation array, both by nanopore sequencing and SoC array. Despite this, fours astrocytomas exhibited co-deletion of *CDKN2A/B* on copy number profiling, thus carrying a far worse prognosis, (CNS WHO grade 4) than the methylation class alone would indicate (likely CNS WHO grade 2). Furthermore, the smear preparation in this case did not reveal any high-grade morphological features, and thus intraoperative identification of the high-grade copy number changes may have altered the surgical approach. Emerging novel molecular biomarkers, such as that of copy number load, can be readily integrated into the analysis and may further aid prognostication of these tumours ^16^. As confidence in these approaches grows, it is likely that traditional morphological methods, such as immunohistochemistry, will become redundant, at least for a subset of cases.

Detection of pathogenic SNVs remains a vital aspect of comprehensive molecular diagnosis. Whilst we demonstrate that next-day detection of SNVs is feasible, it must be acknowledged that panel-based sequencing techniques can achieve much higher coverage of target genes. We achieved an average on-target coverage of approximately 30x using PromethION flow cells, which is improved from the coverage that we achieved with ligation-based MinION approaches. Nevertheless, a minimum coverage of 250x or greater has been recommended for somatic variant detection, and far greater coverage is suggested for detection of low frequency variants ^17^. We propose that whilst nanopore-based SNV detection is a helpful additional data point when securing an integrated diagnosis, it is an adjunct to, and an indication for, conventional panel-based confirmatory sequencing at present and cannot be used, yet, for novel mutation discovery. This is likely to change as further improvements in library preparation, adaptive sampling technology and flow cell performance may offer further gains in on-target coverage.

It must be acknowledged that a common cause of misclassification using nanopore data is due to differences in classifier versions; current nanopore tools have been trained primarily on the original Capper dataset ^6^. A subset of our novel cases was classified using a platform-agnostic tool trained using the latest version of the DKFZ classifier and were able to return a correct diagnosis ^18^. This should prompt the clinical and scientific communities to pool data to generate an intraoperative nanopore classification tool which can incorporate novel entities in the same manner as the current SoC array-based classification tools.

## Conclusion

In conclusion, we demonstrate a novel diagnostic assay which can provide reliable methylation-based tumour classification within a 2-hour intraoperative timeframe, and complete diagnostic molecular profiling, including SVs and SNVs, within 24 hours. We anticipate that this technology has the potential to radically alter the present SoC for brain tumour diagnosis. Therefore, it is vital that concurrent efforts are made to ensure that sufficient healthcare professionals are adequately trained in the analysis of this complex data. The role of the neuropathologist is evolving, as they will be called upon to interpret and decipher multifaceted data in real-time and integrate it with the morphological, clinical and radiological findings. Only by situating this data within the unique clinical context of the patient will better outcomes be achieved, and efficacious tailored therapies be realisable.

The pipeline that we describe is low cost and readily adoptable with a small laboratory footprint. For this technology to be best delivered for the benefit of patients, it is vital that pathways are designed around decentralised, near-patient sequencing. Capital costs of nanopore devices are low and the technique of sequencing can be easily adopted by non-expert laboratories. To leverage maximal reduction in diagnostic turnaround times, only data should move between hospitals; expensive and slow transport of tissue must be reduced to a minimum. It must be acknowledged that a common cause of misclassification using nanopore data is due to differences in classifier versions; a subset of our novel cases were successfully classified using an updated platform-agnostic tool ^18^. This should prompt the clinical and scientific communities to pool data to generate an intraoperative nanopore classification tool which can incorporate novel entities in the same manner as the current SoC array-based classification tools. Efforts must be made to centralise data storage, to ensure reproducible, robust analysis of the data and enable the generation of a nanopore-specific reference cohort.

## Data Availability

De-identified individual participant data that underlie the results reported in this article will be made available upon reasonable request. The ROBIN code used for the analysis is available at https://github.com/looselab/robin.

