## Supplementary Materials for "ROBIN: A unified nanopore-based sequencing assay integrating real-time, intraoperative methylome classification and next-day comprehensive molecular brain tumour profiling for ultra-rapid tumour diagnostics"

### Table of Contents

1. Additional Methods
   i. DNA extraction, quantification and normalisation (Retrospective cohort)
   ii. Library preparation and nanopore sequencing (Retrospective cohort)
   iii. Readfish adaptive sampling
   iv. Nextflow pipeline: integrated methylation classification, SNV and SV calling
   v. Classifier versions
2. Supplementary Figures
   i. Supplementary Data Figure 1: Copy number heatmaps
   ii. Supplementary Data Figure 2: Copy number plot of all astrocytomas in the prospective cohort
   iii. Supplementary Data Figure 3: Pathognomonic fusion events within the intraoperative cohort.
   iv. Supplementary Data Figure 4: Intraoperative sequencing results over 1 hour
3. Supplementary Tables
   i. Supplementary Data Table 1: Overview of Intraoperative cases
   ii. Supplementary Data Table 2: Overview of Retrospective cases
   iii. Supplementary Data Table 3a: Discrepant cases in retrospective cohort
   iv. Supplementary Data Table 3b: Novel cases in retrospective cohort
   v. Supplementary Data Table 4a: Discrepant cases in intraoperative cohort
   vi. Supplementary Data Table 4b: Novel entities in intraoperative cohort
   vii. Supplementary Data Table 5: Change in classification between merged 5mc/5hmc and 5mc only (Retrospective Cohort)
   viii. Supplementary Data Table 6: Summary of selected additional diagnostic information
   ix. Supplementary Data Table 7: MGMT promoter methylation (intraoperative cohort)

**DNA extraction, quantification and normalisation (Retrospective cohort)**

All samples were extracted using the Promega Maxwell® RSC Blood DNA Kit (Catalogue no: AS1400) on a Promega Maxwell® RSC Instrument (Catalogue No: AS4500) using manufacturer's instructions excepting increasing the ligation buffer and proteinase K solutions to 400 µL and 40 µL, respectively. The extraction protocol was based on the methodology stated in the RapindCNS2 supplementary methods [1]. Samples were quantified using a Thermo Scientific NanoDrop™ One Spectrophotometer (Catalogue No: ND-ONE-W) to obtain purity ratios (A260/A280nm and A260/A230 nm) along with DNA concentration (ng/µL). Purity ratio guidance criteria as follows: A260/A280 nm (1.7-1.9) and A260/A230 (>1.9) as determined by the DeepSeq Laboratory. DNA quantification was also measured using an Invitrogen™ Qubit™ 3 Fluorometer (Catalogue No: Q33216) using the High Sensitivity (HS) assay (Catalogue No: Q32854) which is specific for double-stranded DNA. Samples were diluted to 1:10 to bring estimates into the range of the HS kit. Samples >100 ng/µL were diluted to the optimal range using 1x TE buffer (10 mM Tris-HCl and 1 mM EDTA). Samples with a concentration <100 ng/µL were aliquoted neat and prepared as close to the recommended yield as possible (1-2 µg), although samples below this range were still sequenced to ascertain whether the data generated was adequate.

**Library preparation and nanopore sequencing (Retrospective cohort)**

Genomic DNA samples were diluted in nuclease-free water and concentrations were measured using the Qubit 4 Fluorometer (Thermo Fisher Scientific) and the Qubit 1X dsDNA HS Assay Kit (Thermo Fisher Scientific; Q33231). DNA integrity and fragment-length profiles were assessed using the Agilent TapeStation 4200 and the Agilent Genomic DNA ScreenTape Assay (Agilent; 5067-5365 and 5067-5366). To ensure optimum sequencing yields, each genomic DNA sample was sheared using a Covaris g-TUBE (Covaris; 520079). The g-TUBEs were centrifuged at 4,200 rpm, the speed recommended for shearing high molecular weight DNA to ~ 20 kb fragment. Sequencing libraries were prepared from ~ 2 µg of sheared DNA, using the Ligation sequencing DNA V14 protocol (Oxford Nanopore Technologies). This protocol used the Ligation Sequencing Kit (Oxford Nanopore Technologies; SQK-LSK114) and all purification steps were performed using AMPure XP beads (Beckman Coulter; A63882). Each library was run over one MinION flow cell (Oxford Nanopore Technologies; FLO-MIN114 – R10.4.1) on the GridION X5. To maximise sequencing output from the GridION sequencing runs, each flow cell was flushed twice, using the Flow Cell Wash Kit (Oxford Nanopore Technologies; EXP-WSH004) and reloaded with fresh sequencing library.

**Readfish adaptive sampling**

Adaptive, targeted sequencing of the regions-of-interest (ROIs) was performed using the Readfish tool [2] and a BED file of targets. This included 3,487 targets and including every chromosome other than Y as described in Patel et al. [1]. Basecalling for adaptive sampling was carried out on a dedicated Nvidia 4090 GPU running the dorado basecaller using the appropriate FAST mode model and mapping to hg38 with alt chromosomes removed.

**Nextflow pipeline: integrated methylation classification, SNV and SV calling**

Reads were aligned to the GRCh38 reference genome using minimap2 [12] generating BAM files. These sorted, indexed, BAM files were then used as inputs to a new Nextflow pipeline which performs a full molecular characterisation of the tumour sample. This pipeline incorporates several modules from the Oxford Nanopore Technologies 'epi2me-labs' wf-human-variation Nextflow workflow (v.2.3.0) which calls SNPs, structural variants (SV), copy number variations (CNV), aggregates modified base data, and generates reports. Briefly, the packages used for these analyses were Clair3 (v.1.0.8), sniffles2 (v.2.0.7), modkit (github.com/nanoporetech/modkit; v.0.3.0), and QDNASEQ (v.1.34.0)[3–5]. Aggregated modified base data, containing the methylation probabilities, are used to perform tumour classification using three methods: rapidCNS2, Sturgeon (v.0.4.4), and nanoDx (v1.0rc3). Variant calls generated by wf-human-variation (clairS) and those generated separately by clairS_To (v0.1.0), are annotated (ClinVar v.20240708, annovar v.2019-10-24) and reported in an interactive html alignment generated by igv_reports (v1.12.0), highlighting somatic mutations where possible. The methylation of the *MGMT* promoter region is analysed and reported using the method detailed in the rapidCNS2 pipeline. Variants and classifications are reported alongside depth of coverage estimates and a copy number variation plot in a final report (Figure 2). Briefly, the packages used for these analyses were CNVpytor (v.1.3.1), samtools (v.1.13), mosdepth (v.0.3.4), methylartist (v1.2.7), vcftools (v.0.1.16), bedtools (v.2.3.0), annovar with hg38 databases, and R scripts associated with the Rapid-CNS [1] pipeline (Quinlan and Hall, 2010; Wang, Li and Hakonarson, 2010; Danecek et al., 2011; Suvakov et al., 2021; R Core Team, 2022).

**Classifier versions**

Three major brain tumour classifiers have been published and are used in present study: the random-forest based classifier NanoDx [1,6], its subsequent neural-network based implementation, CrossNN [7], and the Sturgeon neural-network based classifier [8]. The Sturgeon classifier is specifically optimised to handle the sparse data typically generated by nanopore sequencing, having been trained using simulated cases generated from sub-sampling methylation array data. On the other hand, CrossNN aims to be a platform-agnostic classifier than can handle both array- and nanopore-derived methylation data. Crucially, all three classifiers are publicly available and are trained using open-access data generated during the development of the antecedent array-based classification[9]. However, it is important to note that the Heidelberg dataset has undergone multiple and substantial subsequent iterations using additional patient data since this initial publication. Newer versions of this reference dataset define novel entities that were not included in earlier classifier versions, and many of these entities are predicated solely on their unique methylation signatures (https://www.molecularneuropathology.org/mnp/classifiers). As such, these nanopore-based classifiers are *a priori* limited in their ability to correctly diagnose these tumour subtypes. Furthermore, methylation data have been shown to have clinical utility in the prognostic stratification of meningiomas, yet such features are absent from the earlier reference data upon which the nanopore classifiers were trained [10,11].

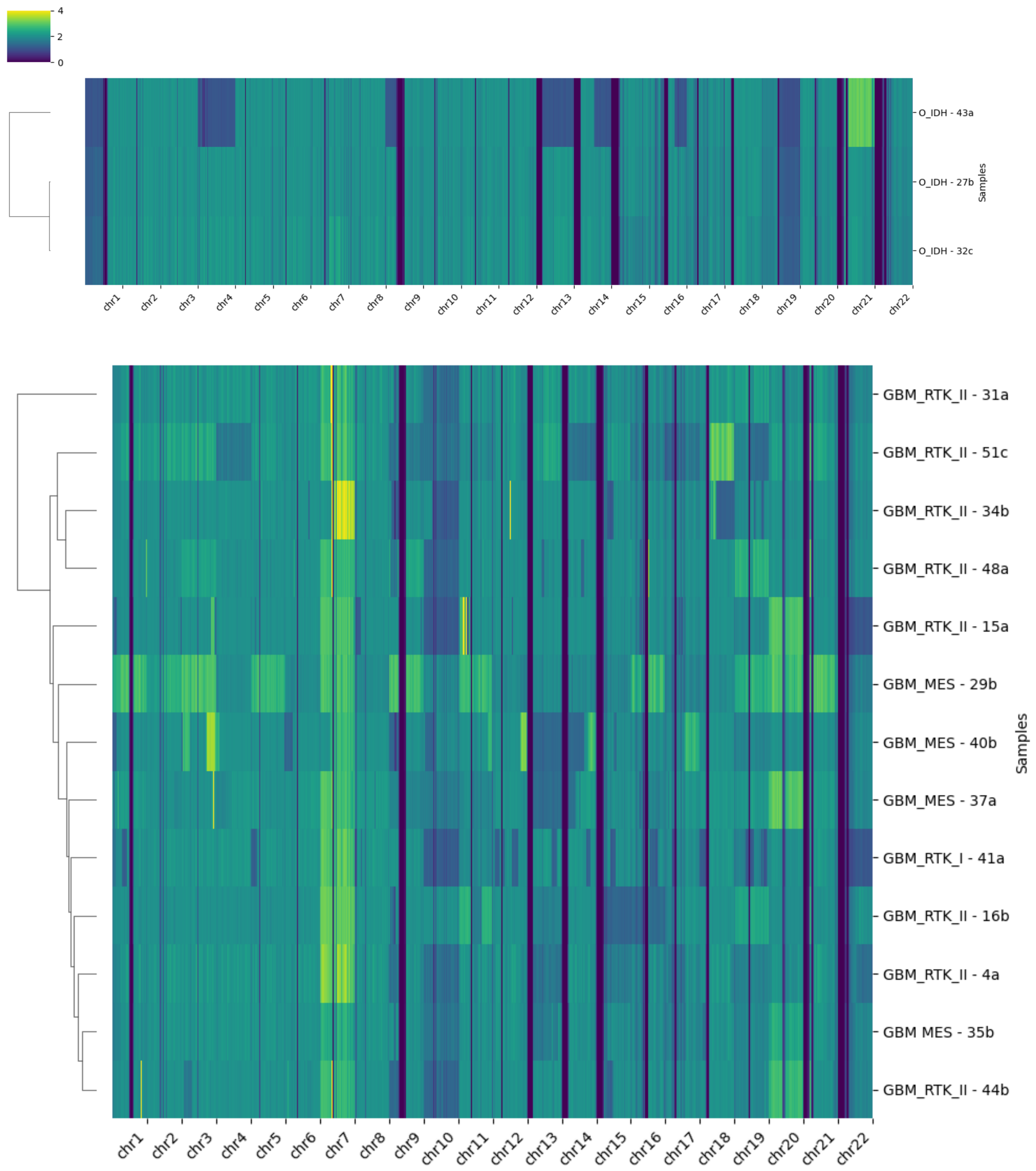


**Supplementary Data Figure 1: Copy number heatmaps**

*Top pane:* Oligodendroglioma exhibiting codeletion of 1p and 19q; *Bottom pane:* Glioblastoma demonstrating gain of chromosome 7 and loss of chromosome 10.

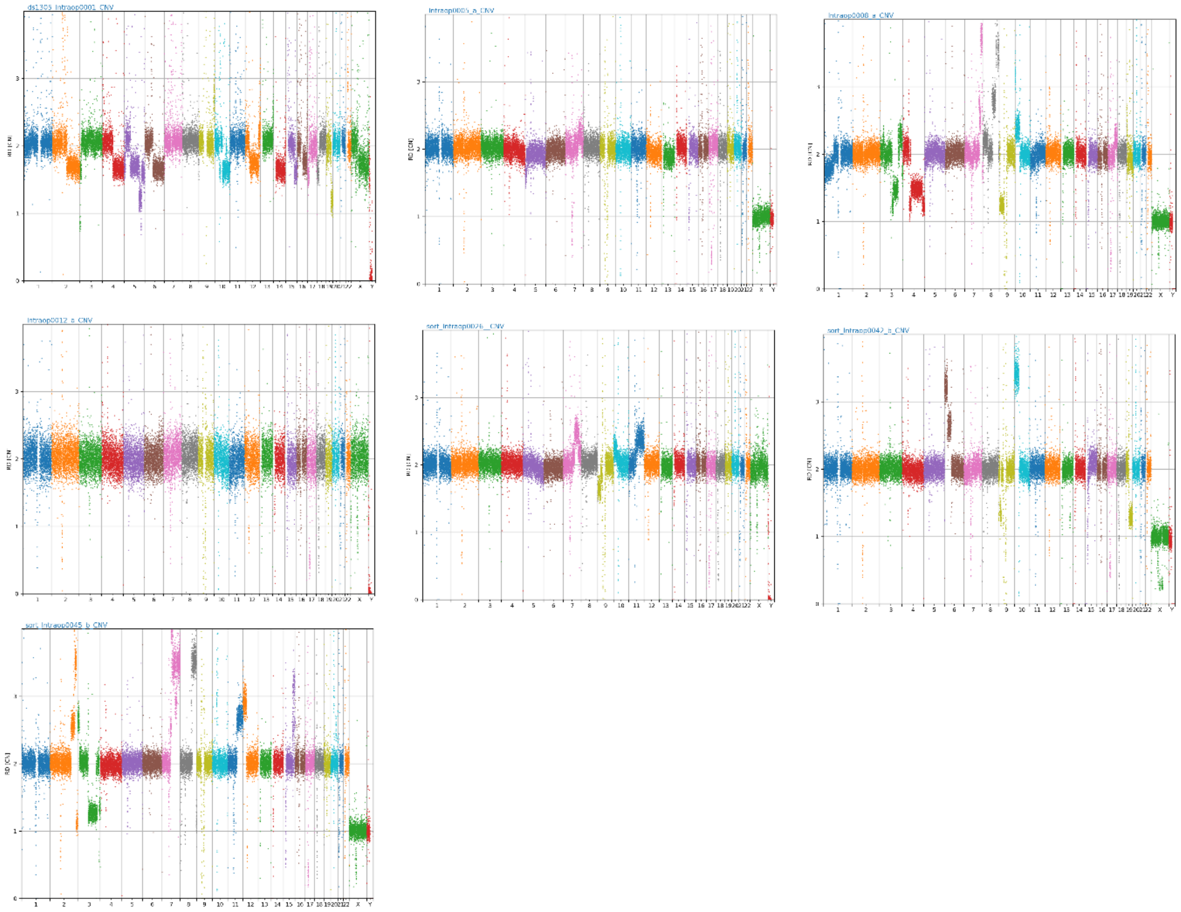

**Supplementary Data Figure 2: Copy number plot of all astrocytomas in the prospective cohort**
Note co-deletion of the CDKN2A/B locus (chr9) in cases 8, 26 and 42. These codeletions were confirmed by SoC testing and have prognostic significance (CNS WHO grade 4), despite all cases classifying on methylation array and nanopore as astrocytoma, lower grade.

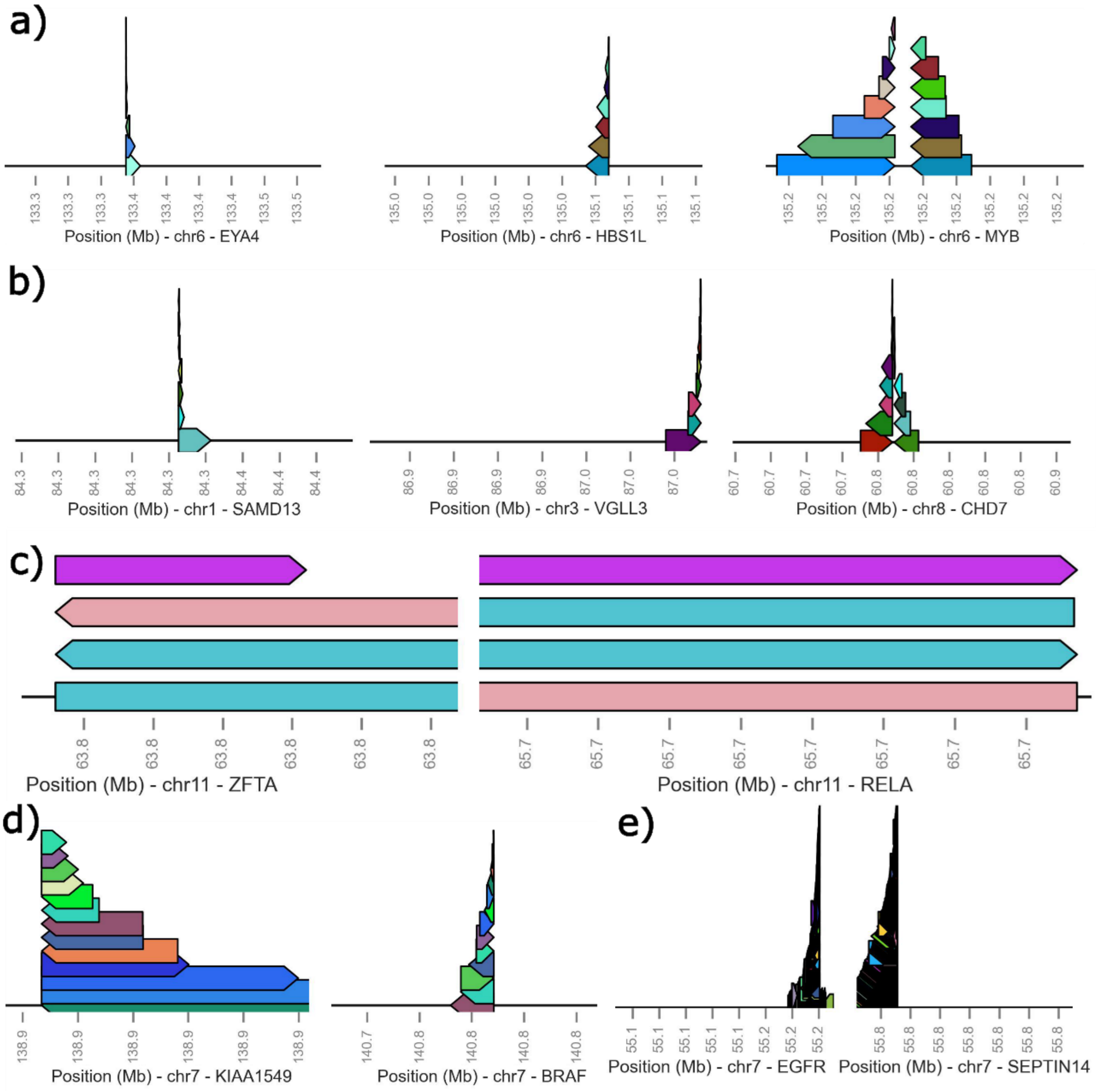


**Supplementary Data Figure 3: Pathognomonic fusion events within the intraoperative cohort.**
Bars each represent a single read, with the colour highlighting the same read mapping across the fusion. A) MYB-altered astrocytoma. B) VGLL-fused intracranial schwannoma. C) ZFTA::RELA fused ependymoma. D) BRAF::KIAA1549 fusion in pilocytic astrocytoma. E) EGFR::SEPTIN14 fused glioblastoma.

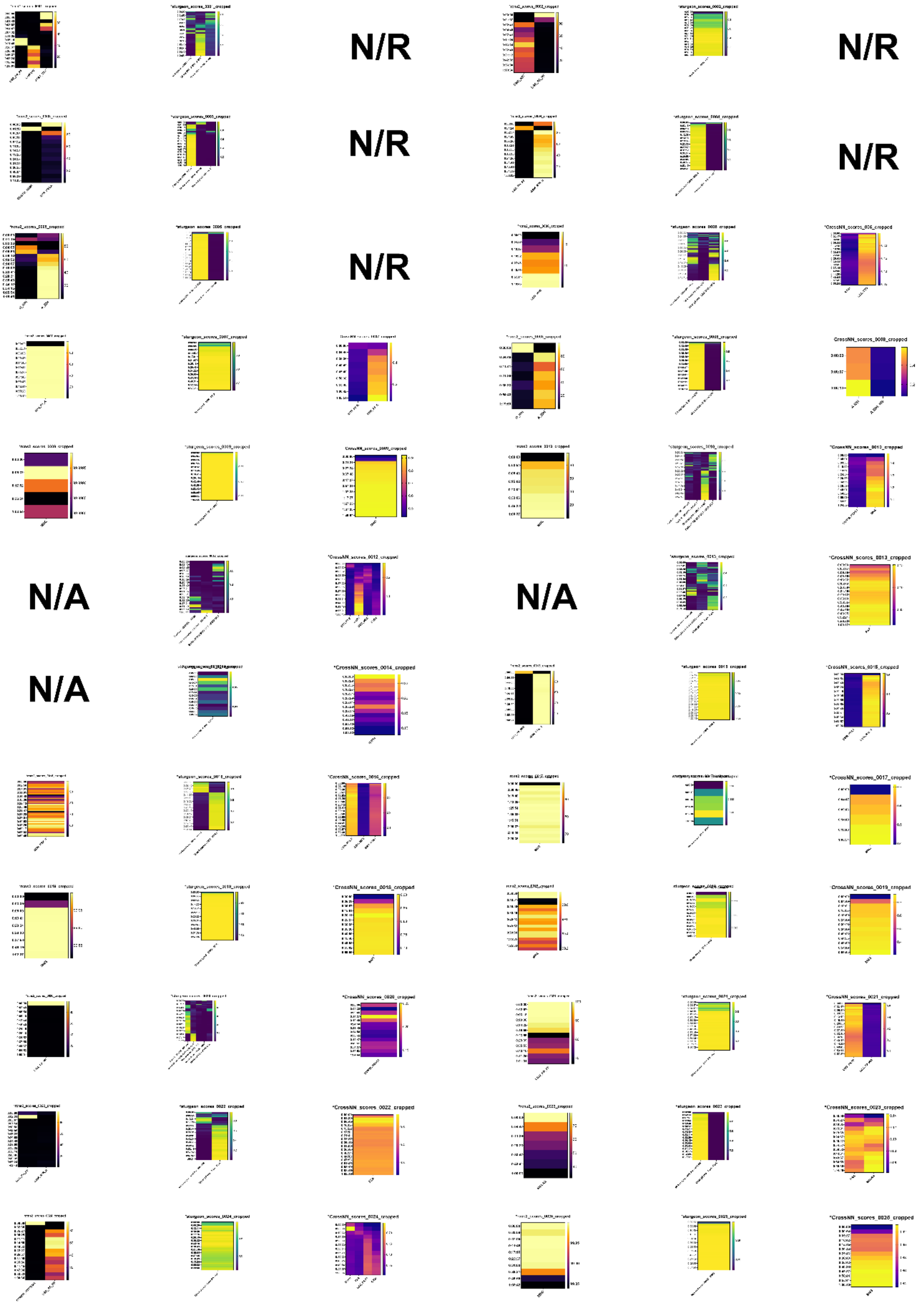


**Supplementary Data Figure 4: Intraoperative sequencing results over 1 hour - Cases 1-25**

Random forest subclasses shown where score reached a threshold of >70 at any timepoint. Sturgeon subclasses shown where score reached a threshold of >0.7 at any timepoint. CrossNN subclasses shown where score reached a threshold of >0.1 at any timepoint. CrossNN was not available for intraoperative reporting for the first 5 cases (N/R). N/A indicates where no subclass met threshold during sequencing.

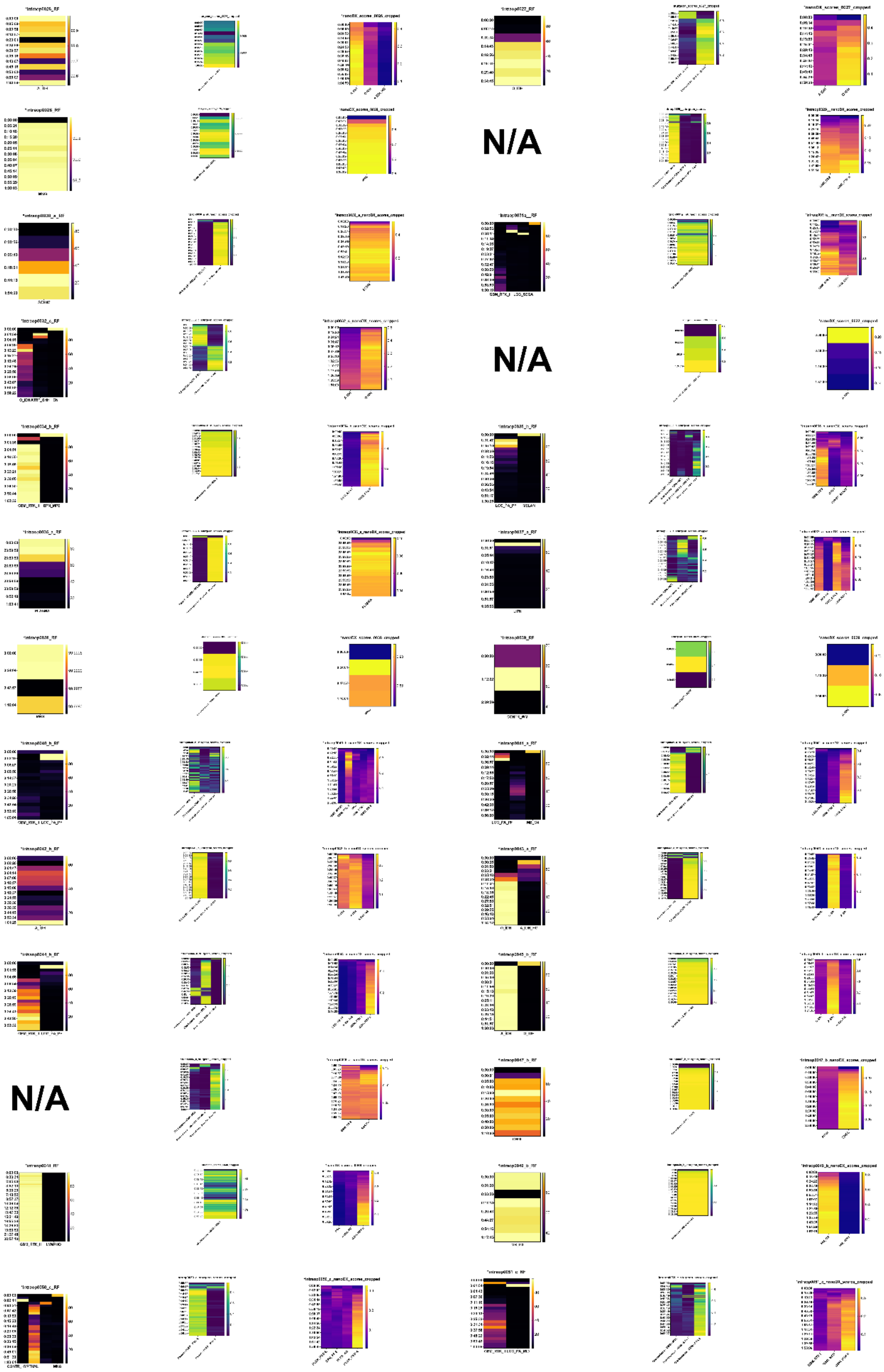


**Supplementary Data Figure 4 (cont.): Intraoperative sequencing results over 1 hour - Cases 26-51**

| SAMPLE | RAPIDCNS2 SUBCLASS | CONF. SCORE | STURGEON SUBCLASS | CONF. SCORE | CROSSNN SUBCLASS | CONF. SCORE | STANDARD OF CARE METHYLATION ARRAY | SCORE | SOC TAT (DAYS) |
|---|---|---|---|---|---|---|---|---|---|
| 1 | A_IDH | 100 | Glioma IDH - A IDH - A IDH | 0.9919 | A IDH | 0.44 | Astrocytoma, IDH-mutant, lower grade | 0.99 | 33 |
| 2 | DMG_K27 | 72 | Glioblastoma - DMG - K27 | 0.9047 | DMG – K27 | 0.33 | Adult-type diffuse high grade glioma, IDH-wildtype, subtypeB | 0.99 | 32 |
| 3 | EPN_RELA | 54.4 | Ependymal - EPN - RELA | 0.9867 | EPN - RELA | 0.79 | Supratentorial ependymoma, ZFTA fusion-positive | 0.99 | 19 |
| 4 | GBM_RTK_II | 100 | Glioblastoma - GBM - RTK II | 0.9636 | GBM, RTK II | 0.48 | Glioblastoma, IDH-wildtype, RTK2 subtype | 0.97 | 33 |
| 5 | A_IDH | 100 | Glioma IDH - A IDH - A IDH | 0.9934 | A IDH | 0.52 | Astrocytoma, IDH-mutant, lower grade | 0.99 | 37 |
| 6 | LGG_MYB | 97.6 | Other glioma - LGG MYB - MYB | 0.9356 | LGG, MYB | 0.31 | Diffuse astrocytoma, MYB or MYBL1-altered, subtype C | 0.99 | 81 |
| 7 | EPN_PF_A | 100 | Ependymal - EPN - PF A | 0.9968 | EPN, PF A | 0.84 | Posterior fossa group A ependymoma, subclass 1c | 0.94 | 29 |
| 8 | A_IDH | 100 | Glioma IDH - A IDH - A IDH | 0.9923 | A IDH | 0.49 | Astrocytoma, IDH-mutant, lower grade | 0.99 | 27 |
| 9 | MNG | 100 | Mesenchymal - MNG - MNG | 0.9996 | MNG | 0.94 | *Not Requested* | - | - |
| 10 | MNG | 100 | Mesenchymal - MNG - MNG | 0.9460 | MNG | 0.54 | Meningioma, benign 1 | 0.99 | 42 |
| 12 | CONTR_HEMI | 5.7 | Glioma IDH - A IDH - A IDH | 0.7563 | A IDH | 0.13 | Astrocytoma, IDH-mutant, lower grade | 0.98 | 39 |
| 13 | PXA | 28 | Other glioma - PXA - PXA | 0.8117 | PXA | 0.17 | Subependymal Giant Cell Astrocytoma | 0.42 | 46 |
| 14 | A_IDH | 25.5 | Glioma IDH - O IDH - O IDH | 0.9095 | O IDH | 0.13 | Diffuse Paediatric-type high grade glioma, RTK1, subclass A | 0.64 | 44 |
| 15 | GBM_RTK_II | 100 | Glioblastoma - GBM - RTK II | 0.9853 | GBM, RTK II | 0.61 | *Not Requested* | - | - |
| 16 | GBM_RTK_II | 99.4 | Glioblastoma - GBM - RTK II | 0.9149 | GBM, RTK II | 0.33 | *Not Requested* | - | - |
| 17 | MNG | 100 | Mesenchymal - MNG - MNG | 0.9981 | MNG | 0.74 | Meningioma, benign 3 | 0.87 | 36 |
| 18 | MNG | 100 | Mesenchymal - MNG - MNG | 0.9996 | MNG | 0.94 | Meningioma, benign 2 | 0.99 | 66 |
| 19 | MNG | 100 | Mesenchymal - MNG - MNG | 0.9983 | MNG | 0.86 | Meningioma, benign 2 | 0.99 | 31 |
| 20 | CONTR_REACT | 42.1 | Control - CONTR - INFLAM | 0.7920 | CONTR, REACT | 0.18 | Chordoma | 0.92 | 34 |
| 21 | LGG_PA_PF | 99.9 | Other glioma - LGG PA - PA | 0.9723 | LGG, PA PF | 0.58 | Infratentorial piloctytic astrocytoma | 0.99 | 27 |
| 22 | GBM_RTK_II | 66.3 | Other glioma - PXA - PXA | 0.9895 | PXA | 0.46 | Pleomorphic xanthoastrocytoma | 0.99 | 29 |
| 23 | MELAN | 60.4 | Melanocytic - MELAN - MELAN | 0.9802 | MELAN | 0.20 | No matching methylation class | <0.3 | 32 |
| 24 | LGG_PA_PF | 77.2 | Glioma IDH - A IDH - A IDH | 0.4802 | LGG, PA PF | 0.19 | Infratentorial piloctytic astrocytoma | 0.51 | 24 |
| 25 | MNG | 100 | Mesenchymal - MNG - MNG | 0.9998 | MNG | 0.95 | Meningioma, benign 3 | 0.47 | 27 |

**Supplementary Data Table 1: Overview of Intraoperative cases 1-25** *Case 11 excluded as metastasis on smear*)

| SAMPLE | RAPIDCNS2 SUBCLASS | CONF. SCORE | STURGEON SUBCLASS | CONF. SCORE | CROSSNN SUBCLASS | CONF. SCORE | STANDARD OF CARE METHYLATION ARRAY | SCORE | SOC TAT (DAYS) |
|---|---|---|---|---|---|---|---|---|---|
| 26 | A_IDH | 100 | Glioma IDH - A IDH - A IDH | 0.9922 | A IDH | 0.50 | Astrocytoma, IDH-mutant; lower grade | 0.99 | 27 |
| 27 | O_IDH | 98.1 | Glioma IDH - O IDH - O IDH | 0.9181 | O IDH | 0.45 | Oligodendroglioma, IDH-mutant and 1p/19q-codeleted | 0.95 | 27 |
| 28 | MNG | 100 | Mesenchymal - MNG - MNG | 0.9995 | MNG | 0.92 | Meningioma, benign 2 | 0.82 | 28 |
| 29 | GBM_RTK_II | 14.8 | Glioblastoma - GBM - MES | 0.9675 | GBM, MES | 0.28 | Glioblastoma, IDH-wildtype, typical mesenchymal type | 0.98 | 36 |
| 30 | SCHW | 90.3 | Nerve - SCHW - SCHW | 0.9586 | SCHW | 0.50 | Schwannoma | 0.52 | 21 |
| 31 | GBM_RTK_II | 69.4 | Glioblastoma - GBM - MES | 0.9453 | GBM, RTK II | 0.41 | Glioblastoma, IDH-wildtype, [typical mesenchymal type] | 0.57 | 32 |
| 32 | O_IDH | 98.5 | Glioma IDH - O IDH - O IDH | 0.9185 | O IDH | 0.49 | Oligodendroglioma, IDH-mutant and 1p/19q-codeleted | 0.94 | 20 |
| 33 | CONTR_HEMI | 7.2 | Glioma IDH - A IDH - A IDH | 0.6111 | CONTR, REACT | 0.08 | Control tissue, white matter (corpus callosum) | 0.93 | 35 |
| 34 | GBM_RTK_II | 100 | Glioblastoma - GBM - RTK II | 0.9180 | GBM, RTK II | 0.50 | Glioblastoma, IDH-wildtype, RTK2 subtype | 0.98 | 18 |
| 35 | PXA | 11.1 | Glioblastoma - GBM - MES | 0.8027 | GBM, MES | 0.28 | Glioblastoma, IDH-wildtype, typical mesenchymal type | 0.99 | 31 |
| 36 | CONTR_REACT | 4.2 | Control - CONTR - INFLAM | 0.9206 | PLASMA | 0.09 | *Not requested (IHC conclusive of Germinoma)* | - | - |
| 37 | GBM_RTK_II | 11.9 | Glioblastoma - GBM - MES | 0.8793 | GBM, RTK II | 0.22 | Glioblastoma, IDH-wildtype, RTK2 subtype | 0.15 | 27 |
| 38 | MNG | 100 | Mesenchymal - MNG - MNG | 0.9997 | MNG | 0.94 | Meningioma, benign 1 | 0.83 | 40 |
| 39 | CONTR_HYPTHAL | 14.8 | Glioma IDH - A IDH - A IDH | 0.7797 | A IDH | 0.06 | control tissue, white matter (corpus callosum) | 0.24 | 42 |
| 40 | GBM_RTK_II | 37.3 | Glioblastoma - GBM - MES | 0.6527 | GBM, RTK I | 0.16 | No matching methylation classes with score >= 0.3 | <0.3 | 34 |
| 41 | GBM_RTK_II | 65 | Glioblastoma - GBM - RTK I | 0.8473 | GBM, RTK I | 0.49 | Glioblastoma, IDH-wildtype, RTK1 subtype | 0.99 | 33 |
| 42 | A_IDH | 99.6 | Glioma IDH - A IDH - A IDH | 0.9877 | A IDH | 0.39 | Astrocytoma, IDH-mutant, lower grade | 0.99 | 33 |
| 43 | O_IDH | 100 | Glioma IDH - O IDH - O IDH | 0.9913 | O IDH | 0.60 | Oligodendroglioma, IDH-mutant and 1p/19q-codeleted | 0.97 | 32 |
| 44 | GBM_RTK_II | 97 | Glioblastoma - GBM - RTK II | 0.9563 | GBM, RTK II | 0.49 | Glioblastoma, IDH-wildtype, RTK2 subtype | 0.98 | 29 |
| 45 | A_IDH | 100 | Glioma IDH - A IDH - A IDH | 0.9928 | A IDH | 0.49 | Astrocytoma, IDH-mutant, lower grade | 0.99 | 29 |
| 46 | LGG_PA_PF | 10 | Other glioma - ANA PA - ANA PA | 0.7444 | GBM, MES | 0.13 | High-grade astrocytoma with piloid features | 0.53 | 27 |
| 47 | CHGL | 71 | Other glioma - CHGL - CHGL | 0.8746 | CHGL | 0.23 | Chordoid glioma, PRKCA mutant | 0.99 | 35 |
| 48 | GBM_RTK_II | 99 | Glioblastoma - GBM - RTK II | 0.8567 | GBM, RTK II | 0.51 | No matching methylation classes with score >= 0.3 | <0.3 | 35 |
| 49 | MB_G3 | 99 | Embryonal - MB G3G4 - G3 | 0.9981 | MB, G3 | 0.82 | Medulloblastoma Group 3, subclass II | 0.99 | 43 |
| 50 | PLEX_PED_A | 77 | Plexus - PLEX - PED A | 0.9935 | PLEX, PED A | 0.39 | Choroid Plexus Papilloma, paediatric subtype | 0.99 | 36 |
| 51 | GBM_RTK_II | 90 | Glioblastoma - GBM - RTK II | 0.8643 | GBM, RTK II | 0.30 | Glioblastoma, RTK1 subtype | 0.79 | 35 |

**Supplementary Data Table 1 Continued: Overview of Intraoperative cases 26-51**

| SAMPLE | RAPIDCNS2 SUBCLASS | CONF. SCORE | STURGEON SUBCLASS | CONF. SCORE | CROSSNN SUBCLASS | CONF SCORE | STANDARD OF CARE METHYLATION ARRAY | CONF SCORE |
|---|---|---|---|---|---|---|---|---|
| 1 | GBM_RTK_II | 88.5 | Glioblastoma - GBM - MES | 0.929 | GBM, RTK II | 0.291 | Glioblastoma, IDH-wildtype, RTK2 subtype | 0.96 |
| 2 | O_IDH | 55.4 | Glioma IDH - O IDH - O IDH | 0.760 | O IDH | 0.272 | NOT REQUIRED | - |
| 3 | MNG | 100 | Mesenchymal - MNG - MNG | 1.000 | MNG | 0.886 | Meningioma, subtype benign, subclass 3 (novel) | 0.91 |
| 4 | LGG_PA_PF | 35.2 | Sella - PITAD STH - STH DNS B | 0.992 | LGG, PA PF | 0.357 | Pilocytic astrocytoma, infratentorial | 0.99 |
| 5 | GBM_RTK_II | 100 | Glioblastoma - GBM - RTK II | 0.850 | GBM, RTK I | 0.274 | Glioblastoma, IDH-wildtype, RTK 1 subtype | 0.99 |
| 6 | EPN_SPINE | 100 | Ependymal - EPN - SPINE | 0.999 | EPN, SPINE | 0.733 | Ependymoma, spinal | 0.99 |
| 7 | O_IDH | 11.8 | Glio-neuronal - DLGNT - DLGNT | 0.985 | O IDH | 0.103 | Diffuse glioneuronal tumour, subtype A (novel) | 0.99 |
| 8 | A_IDH | 95.1 | Glioma IDH - A IDH - A IDH | 0.991 | A IDH | 0.507 | Astrocytoma, IDH-mutant; lower grade | 0.81 |
| 9 | O_IDH | 15.6 | Glio-neuronal - DLGNT - DLGNT | 0.983 | DMG, K27 | 0.115 | Anaplastic neuroepithelial tumour with condensed nuclei | 0.97 |
| 10 | LGG_PA_PF | 96.4 | Sella - PITAD STH - STH DNS B | 0.994 | LGG, PA PF | 0.365 | Midline pilocytic astrocytoma | 0.87 |
| 11 | GBM_RTK_II | 75.6 | Glioblastoma - GBM - RTK II | 0.874 | GBM, RTK II | 0.396 | No matching methylation classes | <0.3 |
| 12 | GBM_RTK_II | 9.9 | Glioblastoma - GBM - MES | 0.985 | GBM, MES | 0.286 | Glioblastoma, IDH-wildtype, mesenchymal subtype | 0.88 |
| 13 | LGG_PA_GG_ST | 17.8 | Other glioma - LGG PA - PA/GG ST | 0.935 | LGG, PA/GG ST | 0.132 | Pilocytic astrocytoma, hemspiheric | 0.99 |
| 14 | PXA | 87.4 | Other glioma - PXA - PXA | 0.993 | PXA | 0.456 | Glioblastoma, IDH-wildtype, [atypical mesenchymal type] | 0.87 |
| 15 | SUBEPN_PF | 94.7 | Ependymal - SUBEPN - ALL | 0.984 | SUBEPN, PF | 0.594 | Posterior fossa subependymoma | 0.99 |
| 16 | O_IDH | 97.1 | Glioma IDH - O IDH - O IDH | 0.954 | O IDH | 0.282 | Oligodendroglioma, IDH-mutant and 1p/19q-codeleted | 0.96 |
| 17 | A_IDH | 75.3 | Glioma IDH - A IDH - A IDH | 0.911 | A IDH | 0.204 | Astrocytoma, IDH-mutant; lower grade | 0.98 |
| 18 | LGG_DNT | 62.6 | Glio-neuronal - DLGNT - DLGNT | 0.844 | LGG, DNT | 0.203 | Dysembryoplastic neuroepithelial tumour | 0.8 |
| 19 | A_IDH | 99.9 | Glioma IDH - A IDH - A IDH | 0.991 | A IDH | 0.549 | Astrocytoma, IDH-mutant; lower grade | 0.99 |
| 20 | EPN_SPINE | 100 | Ependymal - EPN - SPINE | 0.999 | EPN, SPINE | 0.680 | Spinal ependymoma | 0.99 |
| 21 | MNG | 98.9 | Mesenchymal - MNG - MNG | 0.974 | MNG | 0.411 | Meningioma, subclass benign 3 | 0.96 |
| 22 | LGG_PA_PF | 100 | Other glioma - LGG PA - PA | 0.984 | LGG, PA PF | 0.636 | Infratentorial pilocytic astrocytoma | 0.99 |
| 23 | PLEX_PED_A | 89.9 | Plexus - PLEX - PED A | 0.997 | PLEX, PED A | 0.335 | Choroid plexus papilloma, pediatric subtype | 0.99 |
| 24 | SUBEPN_PF | 68.7 | Ependymal - SUBEPN - ALL | 0.816 | SUBEPN, PF | 0.475 | Subependymoma, posterior fossa | 0.99 |
| 25 | CONTR_REACT | 68.5 | Control - CONTR - REACT | 0.817 | CONTR, REACT | 0.166 | MC control tissue, reactive tumour microenvironment | 0.41 |
| 26 | CONTR_CEBM | 67.7 | Control - CONTR - CEBM | 0.974 | CONTR, CEBM | 0.256 | MC High-grade astrocytoma with piloid features | 0.94 |
| 27 | A_IDH | 98.4 | Glioma IDH - A IDH - A IDH | 0.986 | A IDH | 0.503 | MC Astrocytoma, IDH-mutant; lower grade | 0.94 |
| 28 | GBM_G34 | 97.3 | Glioblastoma - GBM - G34 | 0.991 | GBM, G34 | 0.655 | Diffuse hemispheric glioma, H3 G34-mutant | 0.99 |
| 29 | GBM_RTK_II | 78.6 | Glioblastoma - GBM - RTK II | 0.878 | GBM, RTK II | 0.291 | Diffuse paediatric-type high grade glioma, RTK2 subtype, subclass B (novel) | 0.28 |
| 30 | GBM_RTK_II | 99.8 | Glioblastoma - GBM - RTK II | 0.961 | GBM, RTK II | 0.465 | Glioblastoma, IDH-wildtype, RTK2 subtype | 0.35 |

**Supplementary Data Table 2: Overview of Retrospective cases**

| SAMPLE | NANOPORE SUBCLASS | ARRAY SUBCLASS | V12_SCORE | FINAL INTEGRATED DIAGNOSIS | NOTES |
|---|---|---|---|---|---|
| 11 | GBM, RTK II | No matching methylation classes | <0.3 | Glioblastoma, IDH-wildtype, CNS WHO grade 4 | Unclassifiable |
| 14 | PXA | Glioblastoma, IDH-wildtype, [atypical mesenchymal type] | 0.87 | Epithelioid glioblastoma, CNS WHO grade 4, BRAF mutant | Large overlap between PXA and epithelioid GBM |
| 25 | CONTROL REACT | MC control tissue, reactive tumour microenvironment | 0.41 | Diffuse midline glioma, H3 K27-altered, CNS WHO grade 4 | Poor tissue selection |

**Supplementary Data Table 3a: Discrepant cases in retrospective cohort**

| SAMPLE | NANOPORE SUBCLASS | ARRAY SUBCLASS | V12_SCORE | FINAL INTEGRATED DIAGNOSIS | NOTES |
|---|---|---|---|---|---|
| 9 | O_IDH *vs* DMG, K27 vs DLGNT | Anaplastic neuroepithelial tumour with condensed nuclei (novel) | 0.97 | High-grade neuroepithelial tumour, NTRK2 fusion-positive | Novel entity (not included in v11) |
| 26 | CONTROL CEBM | MC High-grade astrocytoma with piloid features | 0.94 | High-grade astrocytoma with piloid features, CNS WHO grade 3 | Novel entity (not included in v11); Poor tissue selection |
| 29 | GBM, RTK II | Diffuse paediatric-type high grade glioma, RTK2 subtype, subclass B (novel) | 0.28 | Glioblastoma, IDH-wildtype, CNS WHO grade 4 (Chr7-/10+) | Novel entity (not included in v11 (low score) |

**Supplementary Data Table 3b: Novel cases in retrospective cohort**

| SAMPLE | NANOPORE SUBCLASS | ARRAY SUBCLASS | V12_SCORE | FINAL INTEGRATED DIAGNOSIS | NOTES |
|---|---|---|---|---|---|
| 12 | CONTROL *vs* A_IDH | Astrocytoma, IDH-mutant, lower grade | 0.98 | Astrocytoma, IDH-mutant, CNS WHO 2 | Low tumour DNA fraction |
| 20 | CONTR_REACT *vs* CHORDOMA* | Chordoma | 0.92 | Conventional Chordoma | Poor tissue selection |
| 22 | GBM_RTK_II *vs* PXA | Pleomorphic xanthoastrocytoma | 0.99 | Pleomorphic xanthoastrocytoma, CNS WHO 3 | Necrosis & microvascular proliferation |
| 24 | LGG_PA_PF *vs* A IDH | Infratentorial piloctytic astrocytoma | 0.51 | Piloctytic astrocytoma, CNS WHO 1 | Low array score |
| 33 | CONTR_HEMI *vs* A IDH | Control tissue, white matter (corpus callosum) | 0.93 | CNS tissue fragments with diffuse infiltration of a glioma | Low tumour DNA purity; post-radiotherapy |
| 35 | PXA *vs* GBM MES | Glioblastoma, IDH-wildtype, typical mesenchymal type | 0.99 | Glioblastoma, CNS WHO 4 | |
| 39 | CONTR_HYPTHAL *vs* A_IDH | control tissue, white matter (corpus callosum) | 0.24 | CNS white matter with focal tumour infiltrates and microvascular proliferation | Low tumour DNA purity |

**Supplementary Data Table 4a: Discrepant cases in intraoperative cohort**

*Sample 20 was discrepant over time during intraoperative sequencing, oscillating between chordoma and control tissue classifications

| SAMPLE | NANOPORE SUBCLASS | ARRAY SUBCLASS | V12_SCORE | FINAL INTEGRATED DIAGNOSIS | NOTES |
|---|---|---|---|---|---|
| 2 | DMG_K27 | Adult-type diffuse high-grade glioma, IDH-wildtype, subtype B | 0.99 | Adult-type diffuse high grade glioma, IDH-wildtype, subtype B, WHO 4 | Novel entity (not included in v11) |
| 13 | PXA | Subependymal Giant Cell Astrocytoma | 0.42 | Glial Neoplasm, NOS | Unclassifiable; Low array score |
| 14 | A_IDH *vs* O IDH | Diffuse Paediatric-type high grade glioma, RTK1, subclass A | 0.64 | Suggestive of Diffuse Paediatric-type high grade glioma, H3 wildtype IDH wildtype CNS WHO 4 | Novel entity (not included in v11); Adult Patient; Low array score |
| 36 | CONTR_REACT *vs* CONTR_INFLAM | *Not performed* | | Germinoma | Novel entity (not included in v11) |
| 46 | LGG_PA_PF *vs* ANA PA | High-grade astrocytoma with piloid features | 0.53 | High grade glioma, methylation profile high grade astrocytoma with piloid features (best corresponding to CNS WHO grade 3) | Novel entity (not included in v11); Low array score |

**Supplementary Data Table 4b: Novel entities in intraoperative cohort**

| RAPIDCNS2_CLASS | RAPIDCNS2_CONF. (%) MERGE 5MC & 5HMC | RAPIDCNS2_CONF. (%) REMOVE 5HMC | CONF. CHANGE |
|---|---|---|---|
| GBM_RTK_II | 88.5 | 91.5 | -3 |
| O_IDH | 55.4 | 59.2 | -3.8 |
| MNG | 100 | 100 | 0 |
| LGG_PA_PF | 33.4 | 31.7 | 1.7 |
| GBM_RTK_II | 100 | 100 | 0 |
| EPN_SPINE | 100 | 100 | 0 |
| O_IDH | 12.5 | - | LGG_PA_PF |
| A_IDH | 94.4 | 60.5 | 33.9 |
| O_IDH | 15.9 | - | GBM_RTK_II |
| LGG_PA_PF | 96.9 | 96.5 | 0.4 |
| GBM_RTK_II | 74 | 96.5 | -22.5 |
| GBM_RTK_II | 10 | 23.2 | -13.2 |
| LGG_PA_GG_ST | 17.4 | - | LGG PA PF |
| PXA | 89.6 | 48.2 | 41.4 |
| SUBEPN_PF | 93.4 | 99.7 | -6.3 |
| O_IDH | 95.6 | - | LGG_DNT |
| A_IDH | 75.3 | - | LGG_DNT |
| LGG_DNT | 66.5 | - | LGG_PA_PF |
| A_IDH | 99.9 | 77.2 | 22.7 |
| EPN_SPINE | 100 | 99.8 | 0.2 |
| MNG | 99.1 | - | SFT_HMPC |
| LGG_PA_PF | 100 | 100 | 0 |
| PLEX_PED_A | 90.3 | 14 | 76.3 |
| SUBEPN_PF | 77.8 | 99.7 | -21.9 |
| CONTR_REACT | 67.8 | 16.5 | 51.3 |
| CONTR_CEBM | 68.2 | - | MB_SHH_INF |
| A_IDH | 98.4 | 81.5 | 16.9 |
| GBM_G34 | 97.5 | 99.5 | -2 |
| GBM_RTK_II | 80.7 | 99.5 | -18.8 |
| GBM_RTK_II | 99.8 | 99.9 | -0.1 |

**Supplementary Data Table 5: Change in classification between merged 5mc/5hmc and 5mc only (Retrospective Cohort)**

Unlike conventional micro-array-based methods, nanopore sequencing can distinguish between both 5mc and 5hmc modifications. For our analysis, 5mC and 5hmC modifications were merged before classification, as input data must maintain concordance with the array-derived reference data upon which the classifiers were trained. However, it is interesting to note that classifier accuracy was significantly impaired if 5hmC data were removed from the analysis, suggesting that these rarer DNA modifications nevertheless provide important diagnostic information. It is conceivable that a nanopore-based reference cohort of suitable size could leverage this additional biological information, further refining the classification of tumour subtypes.

| ASTROCYTOMA | | | | |
|---|---|---|---|---|
| CASE | IDH1 Status | TP53 Status | 1p/19q | CKDN2A/B |
| 1 | p.R132H | p.R81Q | No | Intact |
| 5 | p.R132C | p.R141L | No | Intact |
| 8 | p.R132H | p.R141C | No | Del |
| 12* | *Not found* | *Not found* | No | Intact |
| 26 | p.R132H | p.R141H | No | Del |
| 42 | p.R132H | p.R116W | No | Del |
| 45 | p.R132H | p.A29T | No | Intact |

*Low tumour DNA purity (classified as control tissue)

| GLIOBLASTOMA | | | | |
|---|---|---|---|---|
| CASE | IDH mutation | TERT | TP53 | gain 7/ loss 10 |
| 4 | No | Upstream | p.V65L | Yes (& complex) |
| 15 | No | Upstream | *Not found* | Yes (& complex) |
| 16 | No | Upstream | p.E153K | Yes (& complex) |
| 29 | No | *Not found* | p.R210P | Yes (& complex) |
| 31 | No | Upstream | p.R210Q | Yes |
| 34 | No | Upstream | *Not found* | Yes |
| 35 | No | *Not found* | *Not found* | Yes |
| 37 | No | Upstream | *Not found* | Yes |
| 40 | No | Upstream | *Not found* | Yes (& complex) |
| 41 | No | *Not found* | *Not found* | Yes (& complex) |
| 44 | No | *Not found* | *Not found* | Yes |
| 48 | No | Upstream | *Not found* | Yes |

| OLIGODENDROGLIOMA | | | | |
|---|---|---|---|---|
| CASE | IDH Status | ATRX Status | TERT | 1p/19q codeletion |
| 27 | IDH2 p.R42K | Intact | Upstream | Yes |
| 32 | IDH1 p.R132H | Intact | *Not found* | Yes |
| 43 | IDH1 p.R132H | Intact | Upstream | Yes |

**Supplementary Data Table 6: Summary of selected additional diagnostic information**: *Top pane:* Astrocytoma. *Middle pane*: Glioblastoma. *Lower Pane*: Oligodendroglioma.

| | Nanopore | | | Array | | | |
|---|---|---|---|---|---|---|---|
| Case | MGMT | Methylation | Status (Cutoff = 25%) | Status (Cutoff = 0.3582) | Methylation | Lower CI | Upper CI |
| 1 | 23 | 47.49 | methylated | methylated | 0.9834999 | 0.8424044 | 0.998498 |
| 2 | 31 | 4.99 | unmethylated | unmethylated | 0.0832796 | 0.0189851 | 0.2989573 |
| 3 | 13 | 13.93 | unmethylated | unmethylated | 0.0799639 | 0.0216062 | 0.254882 |
| 4 | 18 | 26.43 | unmethylated | unmethylated | 0.1479952 | 0.0402688 | 0.418302 |
| 5 | 27 | 46 | methylated | methylated | 0.9691314 | 0.8125346 | 0.995622 |
| 6 | 19 | 7.32 | unmethylated | unmethylated | 0.0288208 | 0.0048703 | 0.152502 |
| 7 | 3 | N/A | N/A | unmethylated | 0.0617912 | 0.0128008 | 0.250666 |
| 8 | 35 | 41.38 | methylated | methylated | 0.9257303 | 0.6956579 | 0.985501 |
| 9 | 27 | 7.42 | unmethylated | *Not performed* | | | |
| 10 | 14 | 8.76 | unmethylated | unmethylated | 0.0389551 | 0.0075663 | 0.177297 |
| 12* | 8 | 10.15 | **unmethylated** | **methylated** | 0.6913067 | 0.3825578 | 0.890043 |
| 13 | 11 | 9.49 | unmethylated | unmethylated | 0.0922101 | 0.0264277 | 0.275413 |
| 14 | 29 | 76.21 | methylated | methylated | 0.9967815 | 0.9385104 | 0.999841 |
| 15 | 7 | 60.41 | methylated | *Not performed* | | | |
| 16 | 32 | 60.19 | methylated | *Not performed* | | | |
| 17 | 22 | 6.36 | unmethylated | unmethylated | 0.0543459 | 0.0125038 | 0.206873 |
| 18 | 10 | 6.32 | unmethylated | unmethylated | 0.0289705 | 0.0048849 | 0.1534933 |
| 19 | 14 | 6.94 | unmethylated | unmethylated | 0.0548605 | 0.0127066 | 0.207472 |
| 20 | 20 | 4.84 | unmethylated | unmethylated | 0.047018 | 0.0101 | 0.192622 |
| 21 | 19 | 16.35 | unmethylated | unmethylated | 0.0437162 | 0.0089011 | 0.188769 |
| 22 | 5 | 5.83 | unmethylated | unmethylated | 0.089189 | 0.0244349 | 0.276848 |
| 23 | 9 | 4.21 | unmethylated | unmethylated | 0.0097327 | 0.0010217 | 0.0863 |
| 24 | 18 | 13.63 | unmethylated | unmethylated | 0.0378514 | 0.007396 | 0.171987 |
| 25 | 28 | 11.2 | unmethylated | unmethylated | 0.0237046 | 0.0038069 | 0.13365 |
| 26 | 31 | 31.71 | methylated | methylated | 0.4706067 | 0.0845057 | 0.8954095 |
| 27 | 15 | 19.9 | unmethylated | unmethylated | 0.1315761 | 0.0396036 | 0.357608 |
| 28 | 17 | 13.09 | unmethylated | unmethylated | 0.0982246 | 0.0272641 | 0.297407 |
| 29 | 8 | 10.47 | unmethylated | unmethylated | 0.0172385 | 0.0024562 | 0.1110812 |
| 30 | 25 | 6.89 | unmethylated | unmethylated | 0.0502665 | 0.0110369 | 0.200644 |
| 31 | 4 | N/A | N/A | unmethylated | 0.0915101 | 0.0249663 | 0.2837928 |
| 32 | 19 | 56.19 | methylated | methylated | 0.9971626 | 0.9477443 | 0.999853 |
| 33 | 13 | 21.42 | unmethylated | unmethylated | 0.313453 | 0.1423486 | 0.5567218 |
| 34 | 9 | 19.03 | unmethylated | unmethylated | 0.0553101 | 0.0121695 | 0.2176825 |
| 35 | 8 | 12.13 | unmethylated | unmethylated | 0.0315812 | 0.0058135 | 0.153884 |
| 36 | 10 | 5.12 | unmethylated | *Not performed* | | | |
| 37 | 20 | 5.96 | unmethylated | unmethylated | 0.0333289 | 0.0062487 | 0.15899 |
| 38 | 24 | 13.78 | unmethylated | unmethylated | 0.0185204 | 0.0025313 | 0.123046 |
| 39 | 7 | 17.88 | unmethylated | unmethylated | 0.0682699 | 0.0162367 | 0.245448 |
| 40 | 7 | 8.42 | unmethylated | unmethylated | 0.0152806 | 0.0020423 | 0.105279 |
| 41 | 6 | 19.43 | unmethylated | unmethylated | 0.0197028 | 0.0029762 | 0.119197 |
| 42 | 30 | 42.53 | methylated | methylated | 0.9346444 | 0.7420585 | 0.986129 |
| 43 | 30 | 72.2 | methylated | methylated | 0.9397614 | 0.614964 | 0.99348 |
| 44 | 11 | 22.43 | **unmethylated** | **methylated** | 0.9016779 | 0.4524042 | 0.990272 |
| 45 | 23 | 35.74 | methylated | methylated | 0.9816494 | 0.8421308 | 0.998139 |
| 46 | 26 | 15.06 | unmethylated | unmethylated | 0.3493738 | 0.1657983 | 0.591971 |
| 47 | 24 | 9.41 | unmethylated | unmethylated | 0.0786054 | 0.0198676 | 0.264191 |
| 48 | 6 | 51.63 | methylated | methylated | 0.9839353 | 0.8142391 | 0.998833 |
| 49 | 9 | 8.99 | unmethylated | unmethylated | *Not provided* | | |
| 50 | 5 | 10.43 | unmethylated | unmethylated | 0.2448144 | 0.1004793 | 0.4847506 |
| 51 | 7 | 28.17 | methylated | methylated | 0.9706608 | 0.8207524 | 0.9958341 |

**Supplementary Data Table 7: MGMT promoter methylation (intraoperative cohort)**

*Case 12 suffered from low tumoral DNA fraction
